# Predictors of real-world parents’ acceptance to vaccinate their children against the COVID-19

**DOI:** 10.1101/2021.09.12.21263456

**Authors:** Petros Galanis, Irene Vraka, Olga Siskou, Olympia Konstantakopoulou, Aglaia Katsiroumpa, Ioannis Moisoglou, Daphne Kaitelidou

## Abstract

**Background:** As the COVID-19 pandemic continues to threaten public health, the vaccination of children against the disease appears to be a key factor to control the pandemic.

**Objective:** To investigate the prevalence of parents who have vaccinated their children against the COVID-19 and the factors influencing this decision.

**Methods:** We conducted a web-based cross-sectional study in Greece during the first week of September 2021. The study questionnaire was distributed through social media and a convenience sample was obtained. Only parents with children aged 12-17 years old could participate in the study. We collected socio-demographic data of parents and we measured their attitudes towards vaccination and COVID-19 pandemic.

**Results:** Study population included 656 parents. Mean age of parents was 45.5 years, while most of them were mothers with a high level of education. Regarding vaccination, 27.1% of parents had their children vaccinated against the COVID-19, while almost all children had a complete vaccination history (98.9%). The most important reasons for decline of COVID-19 vaccination were doubts about the safety and effectiveness of COVID-19 vaccines (45.3%) and fear of side effects (36.6%). Regarding the information about the COVID-19 vaccines, parents showed more trust in family doctors than in scientists and the government. Multivariate regression analysis identified that increased parents’ age, increased trust in COVID-19 vaccines, and positive attitude of parents towards vaccination had a positive effect on children’s vaccination.

**Conclusions:** Understanding the factors influencing parents’ decision to vaccinate their children against the COVID-19 is crucial to increase the COVID-19 vaccination coverage rate. Implementation of public health policies is necessary to spread knowledge about COVID-19 vaccines and to regain vaccine confidence.

## Introduction

The Coronavirus disease 2019 (COVID-19) pandemic continues to cause several problems at the individual, social and economic level, although from the beginning of 2021 there are several vaccines with proven safety and efficacy (Baden et al., 2021; Haas et al., 2021; Hall et al., 2021; Polack et al., 2020). Successful management of the pandemic requires vaccination of the general population at a high rate, which may now be as high as 80% (Bartsch et al., 2020). However, the COVID-19 vaccination rate exceeds 80% in only a few studies, and in some cases is extremely low, reaching as low as 28.6% (Galanis et al., 2021a). Until now, COVID-19 vaccine uptake in most countries fall below estimates of the required threshold for herd immunity (Ritchie et al., 2021).

Unfortunately, in recent years there has been an increase in the general population’s negative attitude towards vaccination. Vaccine hesitancy refers to the reluctance of individuals to accept the vaccination offer. According to the World Health Organization, vaccine hesitancy is one of the top ten threats to public health (World Health Organization, 2020). Regarding children’s vaccination, the role of parents is crucial as they decide whether to vaccinate their children. In a recent systematic review, only 50–70% of Canadian 2-year-old children have received all of the recommended vaccines (Schellenberg and Crizzle, 2020). Parental vaccination decisions are complex and a variety of factors influence parents’ decision to vaccinate or not their children such as trust in access to healthcare providers, health perceptions and practices, healthcare system, attitudes toward vaccines, experiences, risks and effects of vaccination, religious issues, trust in science, information sources, emotions, etc. (Caso et al., 2021; Dubé et al., 2018; Nurmi and Harman, 2021; Schellenberg and Crizzle, 2020).

Unfortunately, COVID-19 cases in children continue to increase due mainly to the highly transmissible delta variant (American Academy of Pediatrics, 2021; Tanne, 2021a). It is now clear that parents’ decision to vaccinate their children against the COVID-19 will be critical to control the pandemic. According to a meta-analysis, the overall willingness of parents to vaccinate their children against the COVID-19 is moderate (56.8%) and ranges from 29% to 72.7% (Galanis et al., 2021b). Several factors affect parents’ intention to vaccinate their children against the COVID-19 such as socio-demographic variables, attitudes toward vaccines, knowledge, psychological factors, etc. (Galanis et al., 2021b). However, to the best of our knowledge, no study to date has estimated the proportion of parents who have vaccinated their children against the COVID-19. Thus, the aim of our study was to investigate the prevalence of parents who have vaccinated their children against the COVID-19 and the factors influencing this decision.

## Methods

### Study design and participants

We conducted a web-based cross-sectional study in Greece during the first week of September 2021. From 15 July 2021 until the time of our study, the Greek government offered a free COVID-19 vaccine to all children aged 15-17 years old throughout the country. The vaccine was also offered to children aged 12-14 years old from 30 July 2021. The vaccination was voluntary and was carried out after the agreement of the parents or guardians. Data collection was carried out online using google forms to create an anonymous version of the study questionnaire. The questionnaire was distributed through social media and a convenience sample was obtained. Only parents with children aged 12-17 years old could participate in the study. Information on the purpose and design of the study was provided at the beginning of the on-line questionnaire, and HCWs provided informed consent to participate anonymously in the study. Given that the percentage of children who are vaccinated against the COVID-19 is unknown, we considered a prevalence of 50% to estimate the largest sample size. Thus, considering the precision level as ±5% and the confidence level as 95%, a sample size of 385 parents was obtained. We subsequently decided to substantially increase the sample size to minimize random error. The Ethics Committee of Department of Nursing, National and Kapodistrian University of Athens approved the study protocol (reference number; 370, 02-09-2021).

### Questionnaire

We collected socio-demographic data of parents and we measured their attitudes towards vaccination and COVID-19 pandemic. Regarding socio-demographic data, we collected the following: gender, parents’ age, marital status, educational level, self-perceived financial status, self-perceived health status, parents’ chronic disease, parents’ previous COVID-19 diagnosis, family/friends with previous COVID-19 diagnosis, and living with elderly people or vulnerable groups during the COVID-19 pandemic. We used a five-point Likert scale from 0 (“very poor”), to 4 (“very good”) to measure financial status and self-perceived health status.

Regarding vaccination, we measured children’s and parents’ COVID-19 vaccination uptake, parents’ seasonal influenza vaccination in 2020 and children’s complete vaccination history. In all cases, we used “yes/no” answers. Also, we recorded the reasons why parents refuse to vaccinate their children

Additionally, we recorded attitudes of parents towards vaccination and COVID-19 pandemic by measuring the following variables: self-perceived severity of COVID-19, self-perceived knowledge regarding COVID-19 and COVID-19 vaccines, concerns about the side effects of COVID-19 vaccination, trust in COVID-19 vaccines, the role of vaccination in ensuring public health, and trust in the government, scientists, and family doctors regarding the information about the COVID-19 vaccines. The above variables were measured on a scale from 0 to 10 with higher values indicate greater agreement.

### Statistical analysis

We used numbers (percentages) to present categorical variables and mean (standard deviation) to present continuous variables. The normality of the distribution of the continuous variables was assessed with the Kolmogorov-Smirnov test and normal Q-Q plots. The dependent variable was COVID-19 vaccine uptake in children. When parents vaccinated their children, the dependent variable defined as 1. Socio-demographic variables and parents’ attitudes towards vaccination and COVID-19 pandemic were the independent variables. The relationship between the independent variables and the COVID-19 vaccine uptake in children was assessed with univariate and multivariate logistic regression analysis. Independent variables with p-values <0.20 in univariate logistic regression analysis, were included in multivariate logistic regression analysis to eliminate confounding. We calculated unadjusted and adjusted odds ratios (OR), 95% confidence intervals (CI), and p-values. In multivariate logistic regression model, we applied the backward stepwise model. All tests of statistical significance were two-tailed. Statistical analysis was performed with the Statistical Package for Social Sciences software (IBM Corp. Released 2012. IBM SPSS Statistics for Windows, Version 21.0. Armonk, NY: IBM Corp.).

## Results

Study population included 656 parents. Table 1 presents socio-demographic data of parents. Mean age of parents was 45.5 years, while most of the participants were mothers (75.5%) and married (83.2%). Regarding educational level, 75.8% of parents had a University degree and 41.3% had a MSc/PhD degree. Regarding the COVID-19, 10.2% of parents were diagnosed with the disease and 58.8% of parents had family/friends with a previous COVID-19 diagnosis.

**Table 1.** Socio-demographic characteristics of parents.

| **Characteristics** | **N** | **%** |
| --- | --- | --- |
| Gender |  |  |
| Mothers | 495 | 75.5 |
| Fathers | 161 | 24.5 |
| Parents' age (years)<sup>a</sup> | 45.7 | 7.7 |
| Marital status |  |  |
| Singles | 34 | 5.2 |
| Married | 546 | 83.2 |
| Divorced | 71 | 10.8 |
| Widowed | 5 | 0.8 |
| Educational level |  |  |
| High school | 159 | 24.2 |
| University degree | 497 | 75.8 |
| MSc/PhD degree |  |  |
| No | 385 | 58.7 |
| Yes | 271 | 41.3 |
| Self-perceived financial status |  |  |
| Very poor | 20 | 3.0 |
| Poor | 68 | 10.4 |
| Moderate | 353 | 53.8 |
| Good | 174 | 26.5 |
| Very good | 41 | 6.3 |
| Self-perceived health status |  |  |
| Very poor | 4 | 0.6 |
| Poor | 26 | 4.0 |
| Moderate | 121 | 18.4 |
| Good | 305 | 46.5 |
| Very good | 200 | 30.5 |
| Parents' chronic disease |  |  |
| No | 508 | 77.4 |
| Yes | 148 | 22.6 |
| Previous COVID-19 diagnosis |  |  |
| No | 589 | 89.8 |
| Yes | 67 | 10.2 |
| Family/friends with previous COVID-19 diagnosis |  |  |
| No | 290 | 44.2 |
| Yes | 366 | 55.8 |
| Living with elderly people or vulnerable groups during the COVID-19 pandemic |  |  |
| No | 453 | 69.1 |
| Yes | 203 | 30.9 |

<sup>a</sup> mean, standard deviation

Parents’ attitudes towards vaccination and COVID-19 pandemic are shown in Table 2. Twenty seven point one percent of parents had their children vaccinated against the COVID-19, while almost all children had a complete vaccination history (98.9%). The most important reasons for decline of COVID-19 vaccination were doubts about the safety and effectiveness of COVID-19 vaccines (45.3%), fear of side effects (36.6%), low self-perceived threat of COVID-19 for children (9.5%), and previous children’s COVID-19 diagnosis (6.9%). Most parents were vaccinated against the COVID-19 (82.9%), while the respective percentage for seasonal influenza in 2020 was 54.3%. Parents’ self-perceived knowledge regarding COVID-19 and COVID-19 vaccines was very high, while trust in COVID-19 vaccines was high. Regarding the information about the COVID-19 vaccines, parents showed more trust in family doctors than in scientists and the government.

**Table 2.** Parents’ attitudes towards vaccination and COVID-19 pandemic.

|  | N | % |
| --- | --- | --- |
| Children's COVID-19 vaccination uptake |  |  |
| No | 478 | 72.9 |
| Yes | 178 | 27.1 |
| Parents' COVID-19 vaccination uptake |  |  |
| No | 112 | 17.1 |
| Yes | 544 | 82.9 |
| Parents' seasonal influenza vaccination in 2020 |  |  |
| No | 300 | 45.7 |
| Yes | 356 | 54.3 |
| Children's complete vaccination history |  |  |
| No | 7 | 1.1 |
| Yes | 627 | 98.9 |
| Reasons for decline of parents to vaccinate their children against the COVID-19 |  |  |
| I have doubts about the safety and effectiveness of COVID-19 vaccines | 105 | 45.3 |
| I am afraid of side effects of COVID-19 vaccines | 85 | 36.6 |
| I believe that even if my children get infected with COVID-19, nothing bad will happen to them | 22 | 9.5 |
| My children have already been diagnosed with COVID-19 and the vaccine will not be beneficial for them | 16 | 6.9 |
| Family physician does not allow me to vaccinate my children due to their medical condition | 4 | 1.7 |
| Self-perceived severity of COVID-19<sup>a</sup> | 8.1 | 2.4 |
| Self-perceived knowledge regarding COVID-19<sup>a</sup> | 8.9 | 1.4 |
| Self-perceived knowledge regarding COVID-19 vaccines<sup>a</sup> | 8.6 | 1.8 |
| Concerns about the side effects of COVID-19 vaccination<sup>a</sup> | 6.0 | 3.2 |
| Trust in COVID-19 vaccines<sup>a</sup> | 6.9 | 3.2 |
| COVID-19 vaccination promotes public health<sup>a</sup> | 7.8 | 3.1 |
| Vaccination is beneficial<sup>a</sup> | 8.8 | 2.3 |
| Trust in the government regarding the information about the COVID-19 vaccines<sup>a</sup> | 5.2 | 3.5 |
| Trust in scientists regarding the information about the COVID-19 vaccines<sup>a</sup> | 7.1 | 3.2 |
| Trust in family doctors regarding the information about the COVID-19 vaccines<sup>a</sup> | 8.1 | 2.2 |

<sup>a</sup> mean, standard deviation

Table 3 presents univariate and multivariate logistic regression models. According to univariate analysis, increased parents’ age and higher educational level were associated with children’s COVID-19 vaccine uptake. Moreover, parents without a previous COVID-19 diagnosis and those who were vaccinated against the COVID-19 and seasonal influenza had a greater probability to vaccinate their children against the COVID-19. Increased self-perceived knowledge regarding COVID-19 and COVID-19 vaccines, self-perceived severity of COVID-19, and trust in COVID-19 vaccines, government, scientists and family doctors were related with children’s COVID-19 vaccine uptake. Additionally, positive attitude of parents towards vaccination was associated with increased vaccination of children. On the other hand, concerns about the side effects of COVID-19 vaccination reduced the likelihood of children being vaccinated.

**Table 3.**
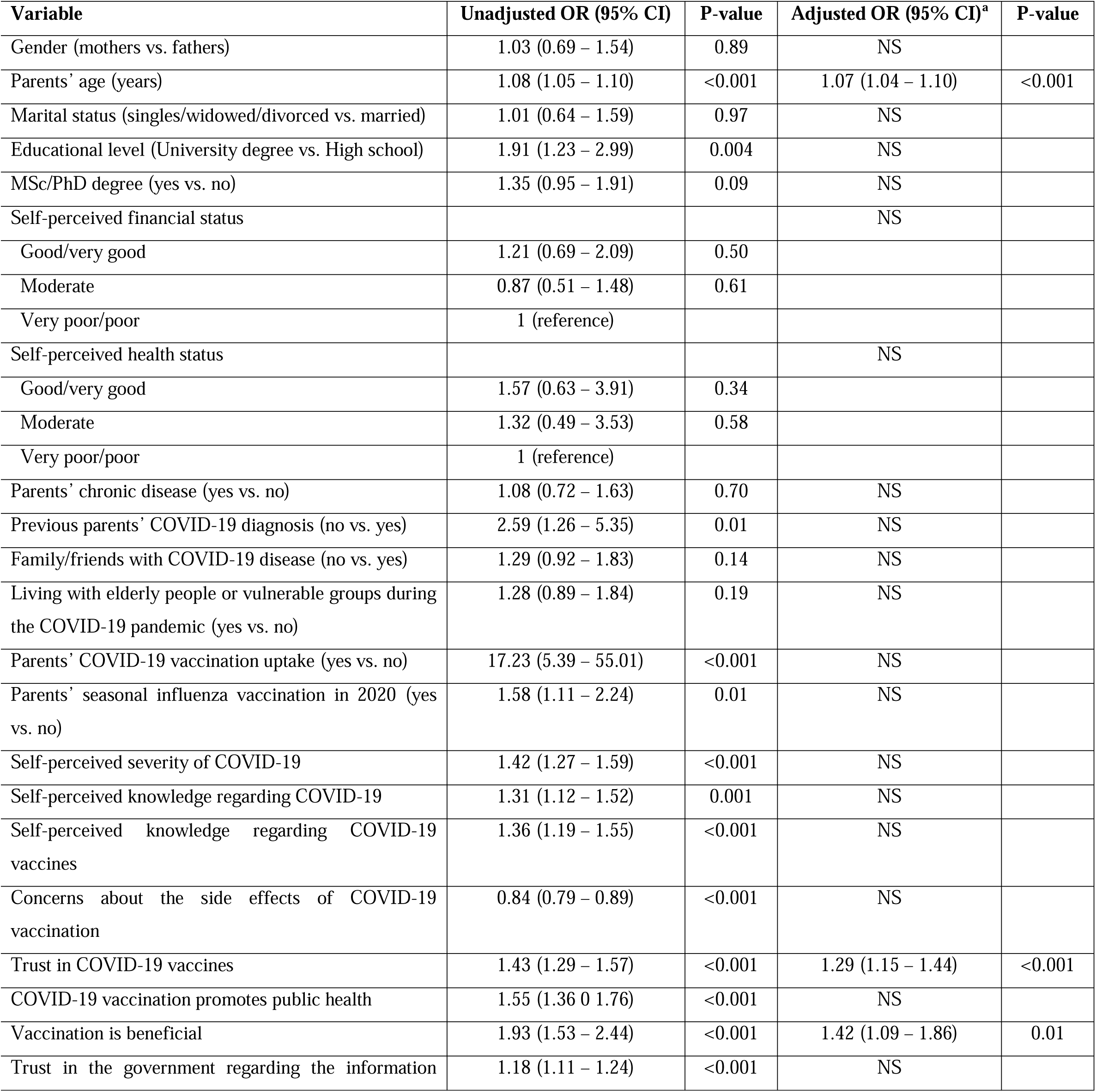
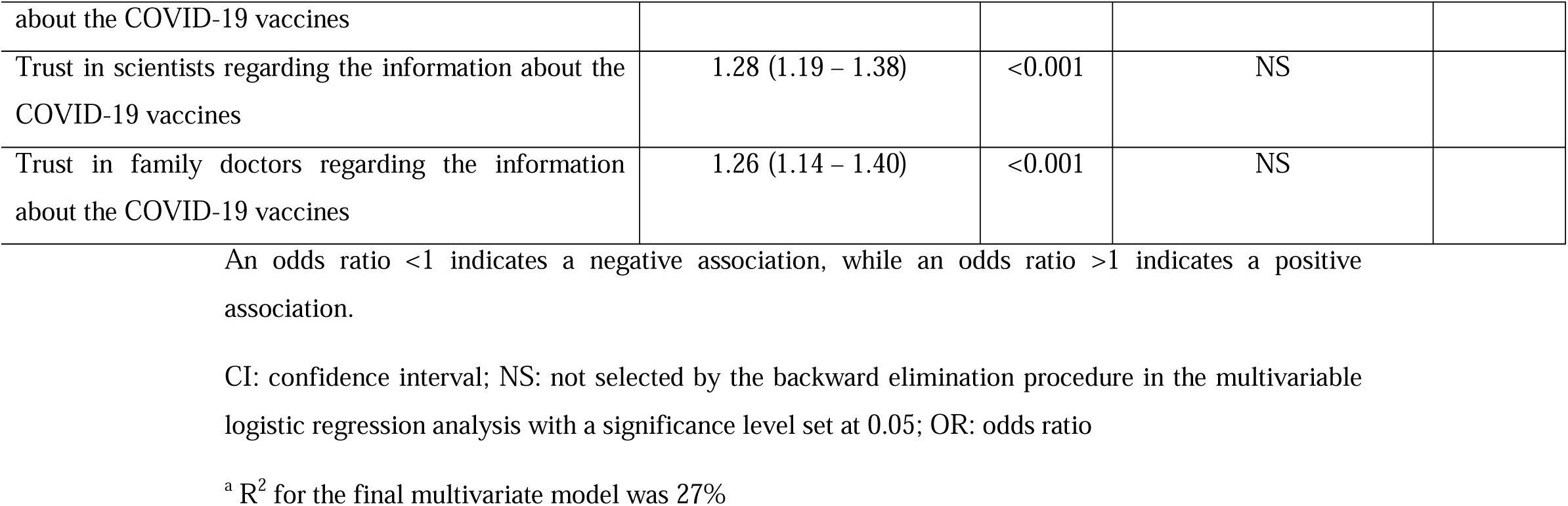
Univariate and multivariate logistic regression analysis with COVID-19 vaccine uptake in children as the dependent variable (reference: COVID-19 vaccine denial).

Multivariate regression analysis identified that increased parents’ age (OR=1.07, 95% CI=1.04-1.10, p-value<0.001), increased trust in COVID-19 vaccines (OR=1.29, 95% CI=1.15-1.44, p-value<0.001) and positive attitude of parents towards vaccination (OR=1.42, 95% CI=1.09-1.86, p-value=0.001) had a positive effect on children’s vaccination.

## Discussion

To our knowledge, this is the first study that estimates COVID-19 vaccine uptake among children and investigates the factors that lead parents to vaccinate their children against the COVID-19. In a sample of parents in Greece, we found that 27.1% of them had already vaccinated their children aged 12-17 years old. This proportion seems low, but the absence of similar studies does not allow us to make direct comparisons with other countries and draw safer conclusions. Furthermore, our study was conducted just one month after the Greek government approved COVID-19 vaccines for children. As evidence from studies increases, it is quite likely that more parents will be persuaded to vaccinate their children against the COVID-19. Early results of randomized controlled trials on the safety and efficacy of COVID-19 vaccines in children are encouraging (Mahase, 2021; Tanne, 2021b), but they are relatively few and it is therefore reasonable for parents to approach COVID-19 vaccines with thoughtfulness and disinclination. Increased parental knowledge about vaccination is a key factor in reducing parents’ concerns and increasing parents’ positive attitude toward vaccination (López et al., 2020; Netfa et al., 2020; Wijayanti et al., 2021).

Consistent with prior studies in adults (Gibbon et al., 2021; Malesza and Bozym, 2021; Schrading et al., 2021; Xu et al., 2021), the most important reasons for decline of COVID-19 vaccination were concerns about the safety and effectiveness of COVID-19 vaccines and fear of side effects. In the case of children, the issue is more complicated, as parents feel responsible for their children’s health and are reluctant to vaccinate their children with a novel vaccine for which there is not yet enough data from studies. It is essential to increase parental knowledge, as this can significantly reduce parents’ vaccine hesitancy (Awad et al., 2019; Jarrett et al., 2015). Parents, even those who are opposed to vaccination, seek as much and more valid scientific information as possible, so that they can make their final decision (Caudal et al., 2020).

An interesting finding of our study is the fact that most parents had been vaccinated against the COVID-19 (82.9%) and more than half had been vaccinated against influenza (54.3%). This finding is extremely encouraging, as it indicates a positive attitude of parents towards vaccination at a time when the COVID-19 vaccination rate among adults is low (Barry et al., 2021; Malesza and Bozym, 2021; Nguyen et al., 2021; The OpenSAFELY Collaborative et al., 2021). Moreover, we found that positive attitude of parents towards vaccination had a positive effect on children’s vaccination. Literature confirms this finding since the successful implementation of childhood vaccination programmes depends to a large extent on the positive attitude of parents (Abdullahi et al., 2016; Alabadi and Aldawood, 2020; Netfa et al., 2020). Awareness campaigns and distribution of leaflets about vaccination have positive effects and are critical in changing the attitudes of parents toward vaccinating their children (Meharry et al., 2014; Wong et al., 2016). Also, individual stories from COVID-19 patients and positive experiences of vaccinated people could promote positive attitude of parents towards vaccination (Kunitoki et al., 2021).

Our multivariate regression model revealed that parents who had more trust in COVID-19 vaccines were more likely to vaccinate their children against the disease. Several studies have already shown that trust in COVID-19 vaccines is related with parents’ willingness to vaccinate their children against the COVID-19 (Skjefte et al., 2021; Yilmaz and Sahin, 2021; Zhang et al., 2020). Vaccine hesitancy today is a major public health issue and is mainly due to a lack of trust. It is essential to develop a fruitful dialogue between citizens, scientists, policymakers and the government, so as to create a relationship of trust and to implement up-to-date health policies on COVID-19 vaccination (Kunitoki et al., 2021). Furthermore, the role of the mass media is particularly important, as the spread of fake news reduces both confidence in COVID-19 vaccines and the intention to vaccinate (Montagni et al., 2021). On-line COVID-19 information from websites is often of low quality and should therefore be systematically monitored by impartial governmental bodies (Cuan-Baltazar et al., 2020; Fan et al., 2020; Joshi et al., 2020). We emphasize the crucial role of family doctors in informing parents as we found that parents trust family doctors more and scientists and government less. For this reason, medical associations should continuously inform and train family doctors on the appropriate way to approach and inform parents on controversial issues such as vaccination.

Our findings should be interpreted in the context of several limitations. Given the convenience sample, the on-line data collection, and the unknown response rate, the extent to which our findings can be generalized to other parents is unknown. For instance, parents with limited internet access were less likely to participate in our study. There may have been an information bias in our study since vaccine uptake was self-reported and some parents may have falsely reported that they vaccinated their children. However, the fact that the questionnaire was completed anonymously may have reduced this bias. Moreover, we have explored several factors that may influence parents’ decision to vaccinate their children, but clearly there are other factors that can be studied such as psychological factors, mass media variables, impact of fake news, etc. Finally, our sample included mainly mothers with a high level of education. Thus, further studies with representative samples are critically needed to draw safer conclusions.

## Conclusions

We provide an early assessment of children’s COVID-19 vaccination status in Greece. Understanding the factors influencing parents’ decision to vaccinate their children against the COVID-19 is crucial to increase the COVID-19 vaccination coverage rate. Future research is needed in this area, as the factors influencing the parents’ decision are miscellaneous. In addition, evidence from studies regarding COVID-19 vaccines is constantly increasing which may influence parents’ attitudes. Mandatory vaccination does not seem to be the most appropriate approach as it generates strong reactions and disagreements. For this reason, the establishment of mutual trust between parents and the scientific community is an essential prerequisite for making a rational decision regarding COVID-19 vaccination in children. Implementation of public health policies is necessary to spread knowledge about COVID-19 vaccines and to regain vaccine confidence.

## Data Availability

Data will be available after reasonable request.

## References

Abdullahi LH, Kagina BM, Cassidy T, et al. (2016) Knowledge, attitudes and practices on adolescent vaccination among adolescents, parents and teachers in Africa: A systematic review. Vaccine 34(34): 3950–3960. DOI: 10.1016/j.vaccine.2016.06.023.

Alabadi M and Aldawood Z (2020) Parents’ Knowledge, Attitude and Perceptions on Childhood Vaccination in Saudi Arabia: A Systematic Literature Review. Vaccines 8(4): E750. DOI: 10.3390/vaccines8040750.

American Academy of Pediatrics (2021) Children and COVID-19: state-level data report. 12 August. Available at: https://www.aap.org/en/pages/2019-novel-coronavirus-covid-19-infections/children-and-covid-19-state-level-data-report/.

Awad S, Abdo N, Yusef D, et al. (2019) Knowledge, attitudes and practices related to influenza illness and vaccination in children: Role of awareness campaigns in changing parents’ attitudes toward influenza vaccination in Jordan. Vaccine 37(25): 3303–3309. DOI: 10.1016/j.vaccine.2019.04.083.

Baden LR, El Sahly HM, Essink B, et al. (2021) Efficacy and Safety of the mRNA-1273 SARS-CoV-2 Vaccine. New England Journal of Medicine 384(5): 403–416. DOI: 10.1056/NEJMoa2035389.

Barry M, Temsah M-H, Aljamaan F, et al. (2021) COVID-19 vaccine uptake among healthcare workers in the fourth country to authorize BNT162b2 during the first month of rollout. preprint, 1 February. Public and Global Health. DOI: 10.1101/2021.01.29.21250749.

Bartsch SM, O’Shea KJ, Ferguson MC, et al. (2020) Vaccine Efficacy Needed for a COVID-19 Coronavirus Vaccine to Prevent or Stop an Epidemic as the Sole Intervention. American Journal of Preventive Medicine 59(4): 493–503. DOI: 10.1016/j.amepre.2020.06.011.

Caso D, Capasso M, Fabbricatore R, et al. (2021) Understanding the psychosocial determinants of Italian parents’ intentions not to vaccinate their children: an extended theory of planned behaviour model. Psychology & Health: 1–21. DOI: 10.1080/08870446.2021.1936522.

Caudal H, Briend-Godet V, Caroff N, et al. (2020) Vaccine distrust: Investigation of the views and attitudes of parents in regard to vaccination of their children. Annales Pharmaceutiques Francaises 78(4): 294–302. DOI: 10.1016/j.pharma.2020.03.003.

Cuan-Baltazar JY, Muñoz-Perez MJ, Robledo-Vega C, et al. (2020) Misinformation of COVID-19 on the Internet: Infodemiology Study. JMIR public health and surveillance 6(2): e18444. DOI: 10.2196/18444.

Dubé E, Gagnon D, MacDonald N, et al. (2018) Underlying factors impacting vaccine hesitancy in high income countries: a review of qualitative studies. Expert Review of Vaccines 17(11): 989–1004. DOI: 10.1080/14760584.2018.1541406.

Fan KS, Ghani SA, Machairas N, et al. (2020) COVID-19 prevention and treatment information on the internet: a systematic analysis and quality assessment. BMJ open 10(9): e040487. DOI: 10.1136/bmjopen-2020-040487.

Galanis P, Vraka I, Siskou O, et al. (2021a) Predictors of COVID-19 vaccination uptake and reasons for decline of vaccination: a systematic review. preprint, 31 July. Public and Global Health. DOI: 10.1101/2021.07.28.21261261.

Galanis P, Vraka I, Siskou O, et al. (2021b) Willingness and influential factors of parents to vaccinate their children against the COVID-19: a systematic review and meta-analysis. preprint, 28 August. Public and Global Health. DOI: 10.1101/2021.08.25.21262586.

Gibbon S, McPhail E, Mills G, et al. (2021) Uptake of COVID-19 vaccination in a medium secure psychiatric hospital population. BJPsych open 7(4): e108. DOI: 10.1192/bjo.2021.924.

Haas EJ, Angulo FJ, McLaughlin JM, et al. (2021) Impact and effectiveness of mRNA BNT162b2 vaccine against SARS-CoV-2 infections and COVID-19 cases, hospitalisations, and deaths following a nationwide vaccination campaign in Israel: an observational study using national surveillance data. The Lancet 397(10287): 1819–1829. DOI: 10.1016/S0140-6736(21)00947-8.

Hall VJ, Foulkes S, Charlett A, et al. (2021) SARS-CoV-2 infection rates of antibody-positive compared with antibody-negative health-care workers in England: a large, multicentre, prospective cohort study (SIREN). Lancet (London, England) 397(10283): 1459–1469. DOI: 10.1016/S0140-6736(21)00675-9.

Jarrett C, Wilson R, O’Leary M, et al. (2015) Strategies for addressing vaccine hesitancy – A systematic review. Vaccine 33(34): 4180–4190. DOI: 10.1016/j.vaccine.2015.04.040.

Joshi A, Kajal F, Bhuyan SS, et al. (2020) Quality of Novel Coronavirus Related Health Information over the Internet: An Evaluation Study. TheScientificWorldJournal 2020: 1562028. DOI: 10.1155/2020/1562028.

Kunitoki K, Funato M, Mitsunami M, et al. (2021) Access to HPV vaccination in Japan: Increasing social trust to regain vaccine confidence. Vaccine: S0264410×21011452. DOI: 10.1016/j.vaccine.2021.08.085.

López N, Garcés-Sánchez M, Panizo MB, et al. (2020) HPV knowledge and vaccine acceptance among European adolescents and their parents: a systematic literature review. Public Health Reviews 41: 10. DOI: 10.1186/s40985-020-00126-5.

Mahase E (2021) Covid-19: Pfizer reports 100% vaccine efficacy in children aged 12 to 15. BMJ (Clinical research ed.) 373: n881. DOI: 10.1136/bmj.n881.

Malesza M and Bozym M (2021) Factors influencing COVID-19 vaccination uptake in an elderly sample in Poland. preprint, 23 March. Public and Global Health. DOI: 10.1101/2021.03.21.21254047.

Meharry PM, Cusson RM, Stiller R, et al. (2014) Maternal Influenza Vaccination: Evaluation of a Patient-Centered Pamphlet Designed to Increase Uptake in Pregnancy. Maternal and Child Health Journal 18(5): 1205–1214. DOI: 10.1007/s10995-013-1352-4.

Montagni I, Ouazzani-Touhami K, Mebarki A, et al. (2021) Acceptance of a Covid-19 vaccine is associated with ability to detect fake news and health literacy. Journal of Public Health (Oxford, England): fdab028. DOI: 10.1093/pubmed/fdab028.

Netfa F, Tashani M, Booy R, et al. (2020) Knowledge, Attitudes and Perceptions of Immigrant Parents Towards Human Papillomavirus (HPV) Vaccination: A Systematic Review. Tropical Medicine and Infectious Disease 5(2): E58. DOI: 10.3390/tropicalmed5020058.

Nguyen L, Joshi AD, Drew DA, et al. (2021) Racial and ethnic differences in COVID-19 vaccine hesitancy and uptake. medRxiv: The Preprint Server for Health Sciences: 2021.02.25.21252402. DOI: 10.1101/2021.02.25.21252402.

Nurmi J and Harman B (2021) Why do parents refuse childhood vaccination? Reasons reported in Finland. Scandinavian Journal of Public Health: 14034948211004324. DOI: 10.1177/14034948211004323.

Polack FP, Thomas SJ, Kitchin N, et al. (2020) Safety and Efficacy of the BNT162b2 mRNA Covid-19 Vaccine. The New England Journal of Medicine 383(27): 2603–2615. DOI: 10.1056/NEJMoa2034577.

Ritchie H, Mathieu E, Rodés-Guirao L, et al. (2021) Coronavirus Pandemic (COVID-19). Available at: https://ourworldindata.org/coronavirus.

Schellenberg N and Crizzle AM (2020) Vaccine hesitancy among parents of preschoolers in Canada: a systematic literature review. Canadian Journal of Public Health 111(4): 562–584. DOI: 10.17269/s41997-020-00390-7.

Schrading WA, Trent SA, Paxton JH, et al. (2021) Vaccination rates and acceptance of SARS-CoV-2 vaccination among U.S. emergency department health care personnel. Academic Emergency Medicine: Official Journal of the Society for Academic Emergency Medicine 28(4): 455–458. DOI: 10.1111/acem.14236.

Skjefte M, Ngirbabul M, Akeju O, et al. (2021) COVID-19 vaccine acceptance among pregnant women and mothers of young children: results of a survey in 16 countries. European Journal of Epidemiology 36(2): 197–211. DOI: 10.1007/s10654-021-00728-6.

Tanne JH (2021a) Covid-19: Cases in children rise sharply in US as doctors call for vaccine approval. BMJ: 2030. DOI: 10.1136/bmj.n2030.

Tanne JH (2021b) Covid-19: FDA authorises Pfizer vaccine for children 12-15. BMJ (Clinical research ed.) 373: n1204. DOI: 10.1136/bmj.n1204.

The OpenSAFELY Collaborative, Curtis HJ, Inglesby P, et al. (2021) Trends and clinical characteristics of COVID-19 vaccine recipients: a federated analysis of 57.9 million patients’ primary care records in situ using OpenSAFELY. preprint, 26 January. Public and Global Health. DOI: 10.1101/2021.01.25.21250356.

Wijayanti KE, Schütze H, MacPhail C, et al. (2021) Parents’ knowledge, beliefs, acceptance and uptake of the HPV vaccine in members of The Association of Southeast Asian Nations (ASEAN): A systematic review of quantitative and qualitative studies. Vaccine 39(17): 2335–2343. DOI: 10.1016/j.vaccine.2021.03.049.

Wong VWY, Lok KYW and Tarrant M (2016) Interventions to increase the uptake of seasonal influenza vaccination among pregnant women: A systematic review. Vaccine 34(1): 20–32. DOI: 10.1016/j.vaccine.2015.11.020.

World Health Organization (2020) Ten threats to global health in 2019. Available at: https://www.who.int/news-room/spotlight/ten-threats-to-global-health-in-2019.

Xu B, Gao X, Zhang X, et al. (2021) Real-World Acceptance of COVID-19 Vaccines among Healthcare Workers in Perinatal Medicine in China. Vaccines 9(7): 704. DOI: 10.3390/vaccines9070704.

Yilmaz M and Sahin M (2021) Parents’ willingness and attitudes concerning the COVID-19 vaccine: A cross-sectional study. INTERNATIONAL JOURNAL OF CLINICAL PRACTICE. DOI: 10.1111/ijcp.14364.

Zhang KC, Fang Y, Cao H, et al. (2020) Parental acceptability of COVID-19 vaccination for children under the age of 18 years: Cross-sectional online survey. JMIR Pediatrics and Parenting 3(2). DOI: 10.2196/24827.

